# Analyzing the amplification of touch sense in visually impaired individuals and comparison with normal individuals

**DOI:** 10.1101/2023.03.10.23287123

**Authors:** Dev Himanshubhai Desai

## Abstract

**Background:** It has been said for ages that if a person loses one of the basic senses like sight or hearing, other senses are amplified to make up for it. This has been substantially proved without a doubt that amplification helps the specially-abled individual in their day-to-day life to an extent. These increases are present but have not been quantified and measured on how much the increases are present.

**Aim:** 1)To estimate and compare sensitivity of touch between visually impaired and normal people and analyze the amplification if present.

**Methodology:** A Cross-Sectional Case-control Study was carried out. Individuals with 6/6 vision and normal in other senses were first examined with the Static Two-point discrimination test to have a baseline value and then different categories of visually impaired individuals were subjected to this test. The test was carried out on the forehead and fingertips of all subjects. Their results were compared using different statistical tools.

**Results:** Total 45 visually impaired individuals with various severity of blindness with 20 Normal individuals taken for the study. Two-point discrimination values for normal individuals (Fingertip=3.43±1.519,Forehead=13.531±2.364), whereas for all and any type of visually impaired individual (fingertip=2.83±0.27,Forehead=13.08±0.26)is statistically(Fingertip, p=0.0101)(Forehead, p=0.4461).R-value for time spent since the disability and degree of amplification is -0.356.

**Conclusion:** A significant difference is present between the values of test between visually impaired and normal individuals. Appreciable that with various degrees of blindness, the average values of two-point-discrimination value are different. Totally blind shows the highest sensitivity.

## Introduction

It is a well-established age-old concept that vision, out of all our senses (seeing, touch, smell, taste, and hearing), is the one that humans utilize the most. It has been the most important instrument in human survival and evolution, nearly overriding all other senses. (1)

The loss of one of the fundamental senses causes a great deal of difficulty for the person, but the loss of eyesight is the most painful. (2) The country has a high rate of blindness, particularly among youngsters. (3) The WHO has established several projects, such as Vision 2020, to reduce the global burden of blindness. (4)

When a person loses one of their primary senses, like as sight or hearing, it is usually assumed that other senses will be heightened to compensate for the loss and restore as much functioning as possible. This is a well-documented truth that virtually everyone is aware of, however there is no formal record on how much the other senses increase when one sense is removed. (5) (6) (7) Many papers and individuals attribute this to a phenomenon known as “Cortical Spasticity” (8). in neurology, which states that when one part of the brain responsible for a particular sense becomes less functional due to the loss of that sense, it will convert to a new sense, giving that new sense more neurons than in other people. This heightens the impression. However, it is also thought that a neuron’s Cortical Spasticity to convert and work for anything else is only true for neurons in its immediate surroundings. (9) (10)

This phenomenon of amplification is well documented for blindness, i.e. loss of vision, in which touch senses amplify to a large extent so that functionality can be restored, (11) (12) (13) but there are few documentations or studies that actually demonstrate HOW MUCH the amplification is, and if it is present, does it correlate to the amount of loss of vision present in the individual, or the length of time the individual has been visually impaired, or both. (14) (15) (16) (17) (18) (13) (19)

To properly assist the individual, it’s crucial to evaluate the degree of amplification existing and how much functioning is returned or not lost. (20) It’s also critical to comprehend the degree of amplification, as Cortical Spasticity is a prevalent explanation, but Center of Vision in occipital lobe(21) and Center of Fine touch where the senses are carried form the body by the Goll and Burdach (22)tract in the dorsal column of the spinal cord ends in Sensory homunculus in parietal Lobe (23)are no way near, yet the auditory senses are getting amplified.

Here, two-point discrimination test (24)was used for the study to determine the fine touch and the sensitivity of the Sensory afferent pathway, Ascending tract and the capacity and sensitivity of sensory homunculus.

## Methodology

A Cross Sectional Case-control Experimental Study was carried and normal individuals and visually impaired individuals were subjected to Static two-point discrimination test under strict supervision to decrease any error.

### Ethical consideration

The research protocol was approved by relevant institutional review boards or ethics committees and that all human participants gave written informed consent, and the authors have no conflict of interest to declare. The Authors have not received any funding from anyone for this research project and have no financial dilemma.

- Inclusion Criteria: -
  - Participants who were willing to take part in the study and willing to give consent
  - Participants who were not suffering from any skin condition or nerve defect
  - For the Case group,
    - Who has been certified as Visually impaired by the Disability agency
  - For the Control group,
    - Participants who have 6/6 vision confirmed by a certified ophthalmologist in the last 4 weeks
- Exclusion Criteria: -
  - Participants not willing to give consent for the experiment
  - Participants who are suffering from any sin condition or any nerve defect
  - Participants with Type 1 diabetes

### Static two-point discrimination test: - (24)

The two point aesthesiometer (25) (26) is take and set at 0(so that both tips are touching each other) and put on the predetermined location on the subject’s body; while putting this points, the subject is asked to call out how may points it can appreciate. The distance between the tips is gradually increased and two tips are simultaneously put on the subject’s body every time and asked how many point are appreciable. This process is repeated till the subject calls out that it can now appreciate two points. At this time, the measurement on the arc measure tape id recorded and the test on that body part is completed. Among diagnostic tests, 2PD assessment is of particular importance because it is often used in clinical practice to evaluate the severity of peripheral nerve injuries and to monitor recovery and response to treatment. (27)

This data was then analyzed using Excel and SPSS and complex advance analysis was carried out trying to simplify the gross amount of data. Averages, Standard deviation (St.dev.), Standard error of Mean(SEM) and Confidence interval 95 (CI95) were various statistical tools used for analysis along with charts and graphs.

## Results

### Discussion

Total of 109 individuals were taken for this prototype study in which 64 were control group with 6/6 vision and the rest 45 were Visually impaired individuals with variable degree of blindness depicted in the certificate of disability awarded to them as it can be seen in Table 1.

**Table 1:** Distribution of Individuals

| Table 1 :- Distribution of Individuals |  |  |  |  |
| --- | --- | --- | --- | --- |
| Total participants in the study :- 60 |  |  |  |  |
| Normal Vision Individuals | All types Of Visually impaired Individuals |  |  |  |
| 64 | 45 |  |  |  |
|  | High Degree of Blindness | Low Degree of Blindness | One eye Blindness | Total Blindness |
|  | 9 | 14 | 2 | 20 |

Table 2 denotes the two-point discrimination value at fingertip. A comparison between normal individuals and all and any category of visually impaired can be seen. Unpaired t-test with a value of p=0.0101 shows that the difference is statistically significant and that the discrimination value at fingertip is much smaller in visually impaired individuals showing their superior sensitivity to discriminate two point and hence their sensory acuity. The findings are depicted in Figure 1 for a better understanding. A lot of studies show that there is an apparent increase in this tactile acuity (12) (13) (19) (28)

**Table 2:** Two point Discrimination value on Fingertips

| Table 2:- Two point Discrimination value on Fingertips |  |  |
| --- | --- | --- |
|  | Normal Individuals | Any and All visually impaired individuals |
| Sample size | 64 | 45 |
| Average | 3.43 | 2.83 |
| Standard deviation | 1.519 | 0.27 |
| p | 0.0101 |  |

**Figure 1:**
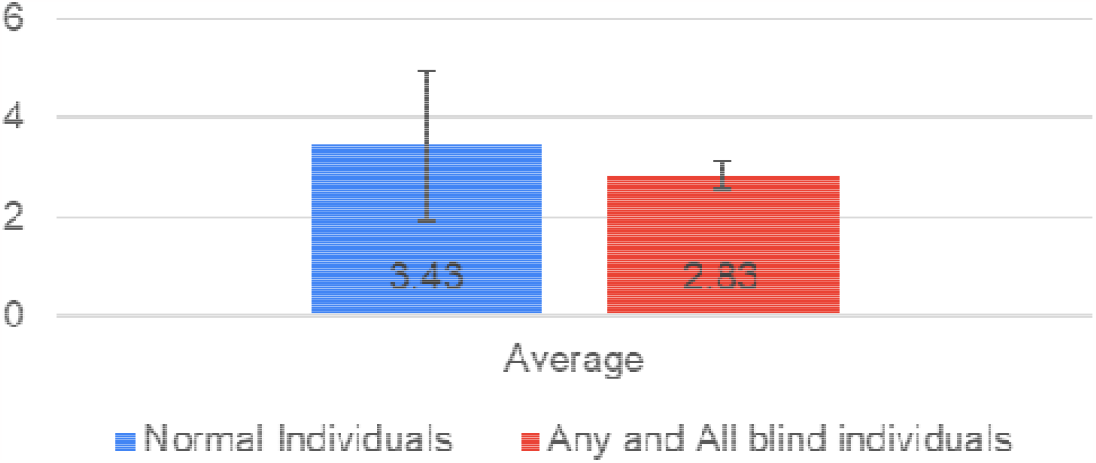
Two point Discrimination value on Fingertip

Similarly, Table 3 and figure 2 here are showing the values of two-point discrimination at forehead. Although very high than fingertip, it is understandable as forehead’s main function is not tactile and touch whereas fingertips’ is. A visible difference can be seen in the values between average discrimination values of Normal individuals and all and any type of visually impaired individuals where the latter has a smaller discrimination value again proving a higher sensitivity.

**Table 3:** Two point Discrimination value on Forehead

| Table 3:- Two point Discrimination value on Forehead |  |  |
| --- | --- | --- |
|  | Normal Individuals | Any and All visually impaired individuals |
| Sample size | 64 | 45 |
| Average | 13.351 | 13.08 |
| Standard deviation | 2.364 | 0.26 |
| p | 0.4461 |  |

**Figure 2:**
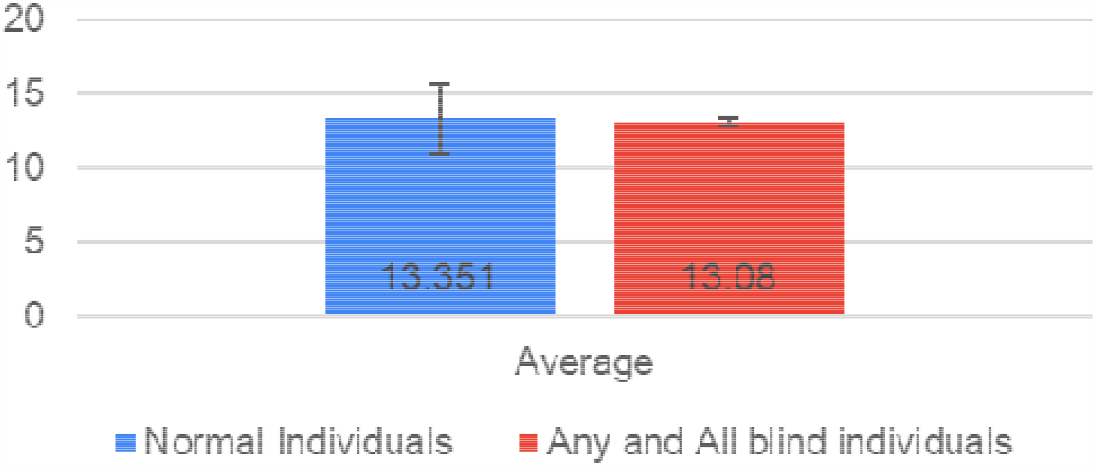
Two point Discrimination value on Forehead

Table 4 and Figure 3 are given here show values of two-point discrimination in individuals of different categories of blindness. Totally blind individuals have the least amount of average discrimination value meaning the highest sensitivity which is followed by individuals with high degree of blindness. Later comes individuals with low degree of blindness and one eye blind individuals. Results of All and any type of individuals will of course be between these different categories as it is simply the average of every individual with any type of blindness i.e. these same individuals from these same categories. The highest amount of discrimination distance meaning the least sensitivity can be seen in normal individuals. Proving that Visually impaired individuals have a higher sensitivity for touch. (29) (13)

**Table 4:** Two point Discrimination value at Fingertip and Forehead in individuals with different degrees of blindness

|  | Fingertip | Forehead |
| --- | --- | --- |
| Normal Individuals | $3.43 \pm 1.519$ | $13.35 \pm 2.364$ |
| All and any type of blindness | $2.83 \pm 0.27$ | $13.08 \pm 0.26$ |
| High degree of Blindness | $2.81 \pm 0.145$ | $13.06 \pm 0.195$ |
| Low degree of blindness | $2.98 \pm 0.222$ | $13.23 \pm 0.268$ |
| One eye Blind | $3.25 \pm 0.070$ | $13.45 \pm 0.079$ |
| Total Blindness | $2.69 \pm 0.257$ | $12.94 \pm 0.216$ |

**Figure 3:**
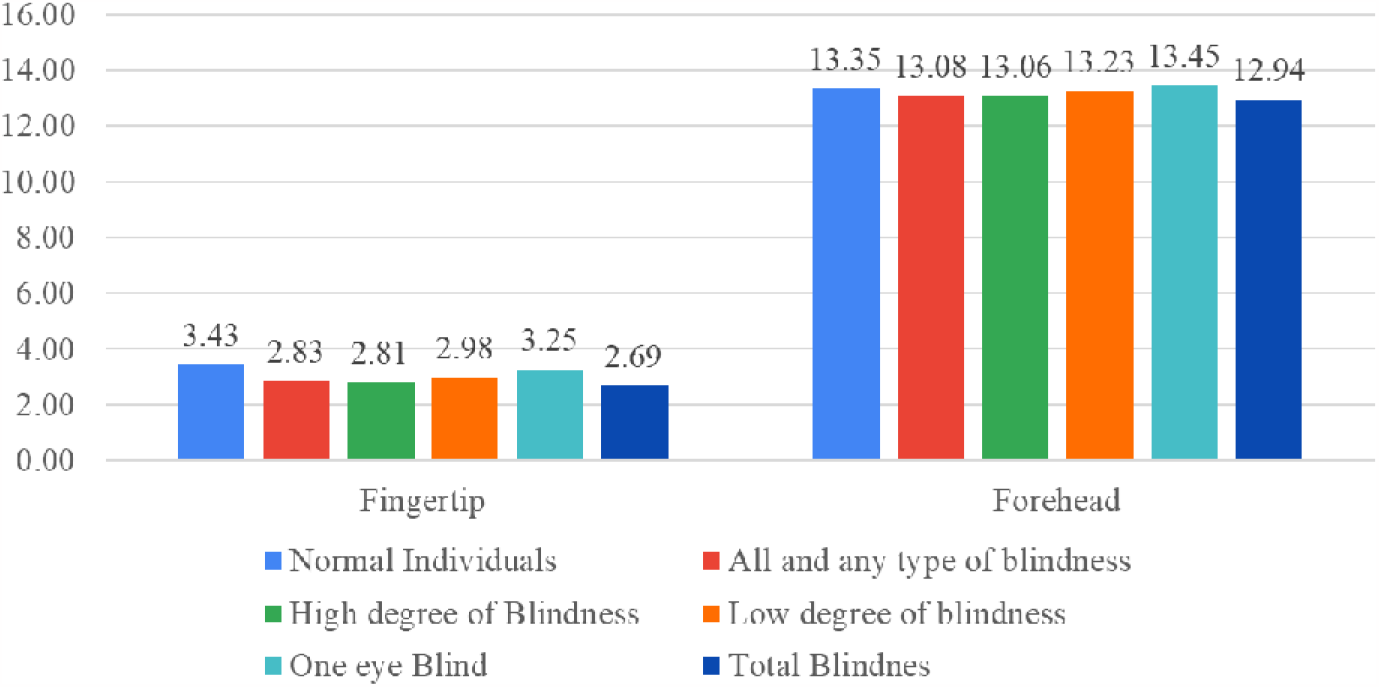
Two point Discrimination value at Fingertip and Forehead in individuals with different degrees of blindnness.

Table 5 shows a very important correlation between the time since the disability is present and its effect on the two-point discrimination value. As for the two-point discrimination value, smaller value means a higher sensitivity, Pearson correlation coefficient in negative actually shows a positive correlation between them and this result is enough to suffice that out of many factors, time spent with disability is an important factor, and that with more time spent with the disability, the amplification of senses is higher. (13) (29)

**Table 5:**
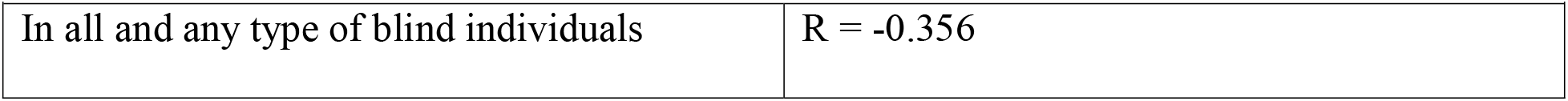
Pearson Correlation between the time passed since the disability and Two-point Discrimination value.

|  |  |
| --- | --- |
| In all and any type of blind individuals | $R = -0.356$ |

## Conclusion

This results are enough to suffice that in visually impaired individuals, a, statistically significant difference in amplification in their sensitivity for touch and sensory acuity is present which is higher than the sensitivity any normal individual possesses and that their two-point discrimination is better. The results are clinically significant and a visible difference is present.

## Data Availability

All data produced in the present study are available upon reasonable request to the authors

## Acknowledgement

The authors would like to thank Blind School Association, Ahmedabad for providing a place with visually impaired individuals to carry on this research. The authors would also like to thank the following individuals for their efforts along with the authors in data collection. We would like to thank Physiology Department of Smt. NHLMMC and Dr. Anita Verma and Dr. Neeraj Mahajan for their constant support. We would like to thank CA Jaladhi Acharya, Dr. Jugal Shah, Dr. Prahasth Dave, Dr. Deval Patel, Dr. Dwija Raval, Dr. Charmi Shah, Dr. Stuti Chauhan, Dr. Tithi Patel, Dr. Yash Chauhan, Dr. Yash Thumar, Dr. Joey Pindaria, Dr. Dev Pandhi, Dr. Jay Patel, Dr. Dev Andharia, Dr. Nishi Shah, Dr. Kshitish Bhramachari and Dr. Tanvi Sahni for their efforts. We would also like to thank Dr. Dholakia (Sanket Speech and Hearing Clinic) for his help in the development process of the methodology.

## Notes

### Competing Interest Statement

The authors have declared no competing interest.

### Funding Statement

This study did not receive any funding

### Author Declarations

IRB of Smt. NHLMMC (Shrimati Nathiba Hargivondas Lakhmichand Municipal Medical College) gave Ethical Approval for this work.

